# Leveraging Open-Source Large Language Models to Identify Undiagnosed Patients with Rare Genetic Aortopathies

**DOI:** 10.1101/2025.09.05.25333227

**Authors:** Pankhuri Singhal, Zilinghan Li, Ze Yang, Tarak Nandi, Colleen Morse, Zachary Rodriguez, Alex Rodriguez, Volodymyr Kindratenko, Giorgio Sirugo, Reed E. Pyeritz, Theodore Drivas, Ravi Madduri, Anurag Verma

**Affiliations:** Department of Medicine, Division of Translational Medicine and Human Genetics, University of Pennsylvania Perelman School of Medicine, Philadelphia, PA, 19104, USA; Data Science and Learning Division, Argonne National Laboratory, Lemont, IL, 60439, USA; The Grainger College of Engineering, Department of Electrical and Computer Engineering, University of Illinois Urbana-Champaign, Urbana, IL, 61801, USA; National Center for Supercomputing Applications, University of Illinois Urbana-Champaign, Urbana, IL, 61801, USA; Consortium for Advanced Science and Engineering, The University of Chicago, Chicago, IL, 60637, USA

## Abstract

Rare genetic aortopathies are frequently undiagnosed due to phenotypic heterogeneity, and delayed diagnosis can lead to fatal cardiac outcomes. While genetic testing can enable early proactive interventions, it relies on primary care physicians to recognize a genetic basis for symptoms and then refer patients to clinical genetics. Broad-scale screening methods are needed to identify cases that do not fit an obvious diagnostic pattern. Clinical notes, rich in narrative details, may support the automated flagging of patients for genetic testing. Given the strength of Large Language Models (LLMs) in processing unstructured text, we developed an open-source LLM-enabled genetic testing recommendation pipeline, which leverages retrieval augmented generation (RAG) on curated genetic aortopathy-related corpora to utilize relevant clinical knowledge for identifying patients likely to benefit from genetic testing. The pipeline was validated using 22,510 patient progress notes from 500 individuals (250 cases, 250 controls) in the Penn Medicine BioBank, and successfully categorized 425 out of 499 patients, with one case requiring further clinician evaluation due to incomplete information. The pipeline achieved a patient-level recommendation accuracy of 0.834, precision of 0.835, sensitivity of 0.831, specificity of 0.836, F1-score of 0.833, and F3-score of 0.832. Our LLM-enabled workflow integrating RAG showed strong performance in recommending genetic testing for patients with rare genetic aortopathies. These findings illustrate the feasibility of using open-source LLMs to support identification of patients who may benefit from genetic testing based on free-text clinical notes, providing a potential decision-support tool to assist clinicians in earlier recognition of rare genetic disease risks.

## Introduction

Rare genetic aortopathies, such as Marfan syndrome and Loeys-Dietz syndrome, are inherited connective tissue disorders that weaken the walls of the aorta, increasing the risk of life-threatening complications like aneurysm or dissection. These conditions are frequently undiagnosed until a major cardiac event, such as aortic dissection, occurs.^1–3^ The heterogeneity of symptoms across the affected patient population makes timely diagnosis a challenge.^4^ Many adults with genetic disorders are reported to have taken more than five years from experiencing initial symptoms to receiving a genetic disease diagnosis,^3^ and these rates are likely even higher for rare genetic conditions. Early screening of at-risk individuals can enable timely medical management and surgical prophylaxis to improve survival rates. Currently, screening methodologies rely on specialist referral to clinical genetics, manual chart review, and primary care physicians’ recognition of subtle phenotypic patterns.^5^ The lack of adaptable tools for broad-scale flagging of patients across healthcare settings has resulted in the underdiagnosis of affected patients.

Free-text clinical notes contain observations and narratives regarding a patient’s health history that can provide evidence to support recommendations for genetic testing. Large language models (LLMs) have shown promise in extracting clinically relevant insights from these unstructured notes.^6–9^ Additionally, unlike encoder-only models such as BERT and Med-BERT,^10, 11^ which are mostly suitable for classification-only tasks, LLMs offer generative capabilities that enable the generation of reasoning for their outputs, potentially enhancing clinical decision-making. While proprietary models like OpenAI’s GPT series demonstrate strong performance across various domains, including medical question answering, their limited adaptability and deployability pose challenges for integration into healthcare systems. Critically, these proprietary models often require data to be sent outside healthcare system firewalls, creating significant privacy concerns in sensitive healthcare environments. Open-source LLMs, when tailored with domain-specific medical knowledge and optimized for local deployment, offer a compelling alternative that supports both scalability and data privacy. This is particularly crucial for rare disease screening in closed and secure clinical infrastructure focused on preserving patient privacy and ensuring computational efficiency.

In this work, we developed an end-to-end pipeline designed for deployment using open-source LLMs within secure clinical environments. Using rare genetic aortopathies as a test case, we evaluate whether LLMs can support identification of patients who may benefit from genetic testing based on clinical narrative data.

## Results

### Evaluation framework and model configuration

We evaluated our pipeline on a dataset of 500 patients (250 cases and 250 controls) with 22,510 clinical notes in total within the Penn Medicine’s health system. Each patient’s record comprised a variable number of notes, reflecting diverse clinical patterns across individuals. The case cohort consisted of patients who were referred for genetic testing by a physician; for these patients, only notes from the **12 months prior to the testing order date** were included. The control cohort comprised age- and sex-matched patients, for whom all available clinical notes were used for analysis.

Our pipeline generates a recommendation for genetic testing based on the contents of each note. The primary objective was to minimize missed diagnoses (false negatives), a particularly critical task in rare disease detection. Thus, we selected the F3-score as our primary metric to evaluate performance, prioritizing sensitivity over precision. Llama 3.1-8B-Instruct^12^, an open-source LLM from Meta, was used as the base model for the recommendation pipeline, with all experiments conducted at an LLM temperature of 0.7 to balance response flexibility and reliability. Both the model and temperature were selected based on performance on a development cohort of 100 patients (50 cases and 50 controls). Additionally, we evaluated the confidence of the LLM recommendation based on the probabilities of the LLM outputs. If the confidence for a given clinical note fell below a threshold of 0.5, a retrieval augmented generation (RAG)^13^ module was automatically triggered to retrieve relevant clinical information from curated aortopathy-related literature, augment the additional information with the clinical notes, and generate a refined prediction.

### Note-level performance: fragmented context limits diagnostic precision

Given the temporal nature of clinical notes data, the pipeline first generates a recommendation evaluation for each note, while accounting for notes that may not be relevant to rare disease symptoms. At the note level, the pipeline achieved an accuracy of 0.753, sensitivity of 0.630, precision of 0.242, F1-score of 0.350, and F3-score of 0.543 (Figure 1a-b). 500 patients were comprised of 22,510 notes, of which 20,720 (92.05%) successfully generated valid recommendations, while 1,790 (7.95%) failed to generate the output in the desired format for result parsing and were therefore excluded from further analysis.

**Figure 1.**
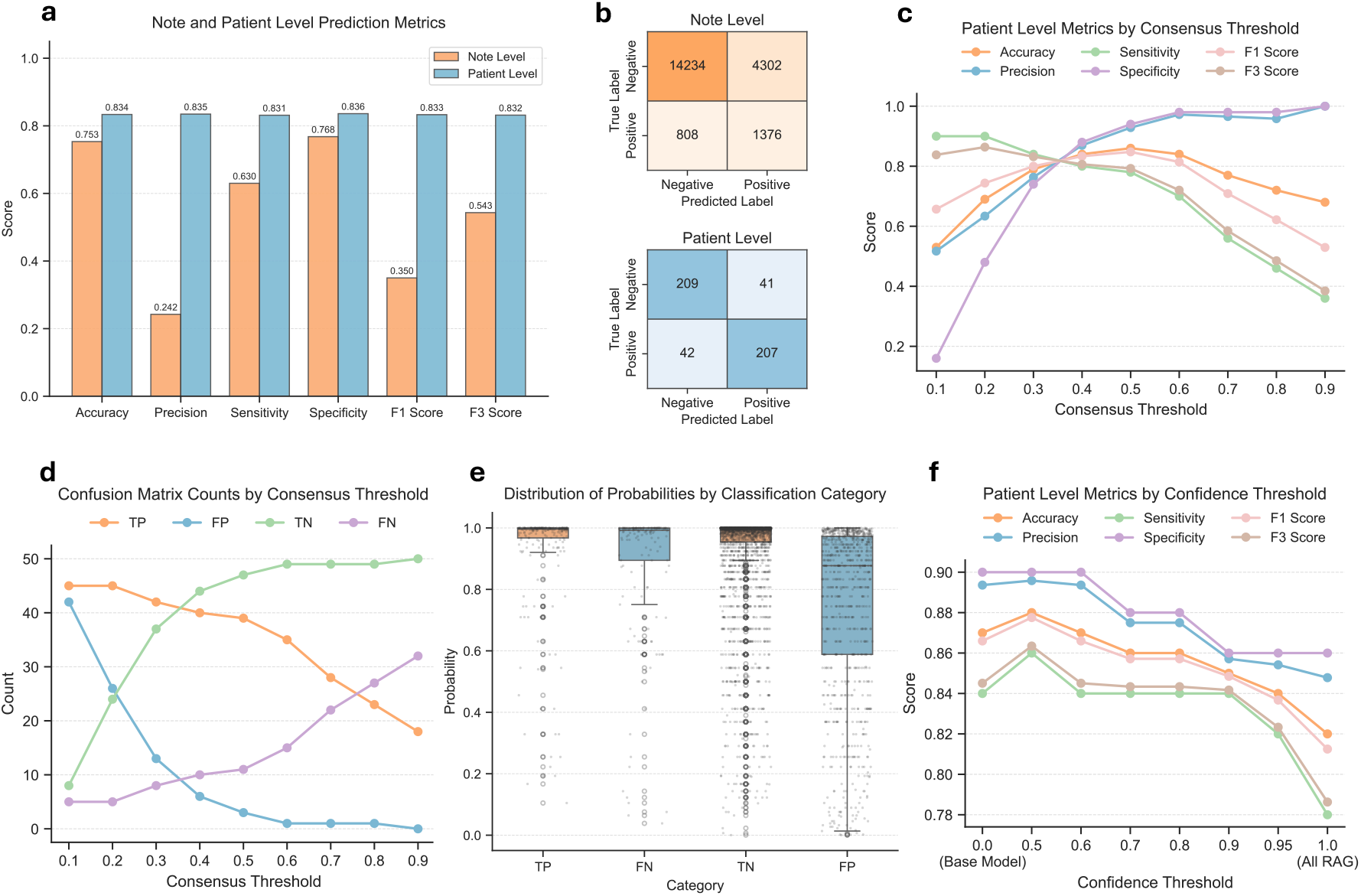
Overview of the pipeline performance. **a** Note and patient-level performance of the pipeline with an optimal confidence threshold of 0.5 and an optimal consensus threshold of 0.4 on the testing dataset (250 cases and 250 controls). **b** Note and patient-level confusion matrices for the pipeline on the testing dataset with optimal threshold settings. **c** Patient-level performance of the pipeline across varying consensus thresholds for note-level recommendation aggregation on a development dataset (50 cases and 50 controls). **d** Patient-level confusion matrix counts across varying consensus thresholds on the development dataset, illustrating the trade-off between sensitivity and specificity as the consensus requirement increases. **e** Distribution of the LLM-generated output probabilities across different classification categories on the patient notes from a development dataset (50 cases and 50 controls, and each patient have various numbers of notes), shown as box plots with individual note-level observations overlaid. **f** Performance comparison of the base model-only, RAG-augmented pipeline with various confidence thresholds, and all-RAG run on the development dataset.

Note-level performance showed relatively high false positive and false negative rates, which was expected as isolated notes may lack sufficient context for comprehensive diagnosis, and not every note contains information relevant to aortopathy. Beyond applying a time-based filter that retains clinical notes within 12 months prior to the genetic testing order date to prevent information leakage, no additional preprocessing filters were used when selecting notes; all notes were included in the analysis. Consequently, some notes are expected to be sparse or clinically irrelevant. Diversity in note relevance may arise depending on the clinician’s style of writing clinical notes, preset templates offered in the physician portal may vary by department or health system, and some notes may be autogenerated by an administrative action, such as ordering a lab test, without inclusion of other relevant clinical information pertaining to the patient. Collectively, these factors limit diagnostic precision when evaluating recommendation performance at the note level.

### Patient-level performance: longitudinal aggregation improves diagnostic accuracy

A final recommendation for each patient was determined by aggregating predictions across all notes per patient using a consensus-based aggregation strategy, in which a patient was flagged if the proportion of notes recommending genetic testing exceeded a consensus threshold. To determine the optimal consensus threshold, patient-level model performance was evaluated across a range of consensus thresholds from 0.1 to 0.9 on a randomly sampled development dataset (50 cases and 50 controls), with the results shown in Figure 1c-d. Performance metrics varied substantially across this range, reflecting the expected sensitivity-specificity tradeoff. At a threshold of 0.4, the model achieved a sensitivity of 0.800, specificity of 0.880, precision of 0.870, F1-score of 0.833, and F3-score of 0.807. Increasing the threshold to 0.5 improved specificity to 0.940 and precision to 0.929, but at the cost of a reduction in sensitivity to 0.780 and F3 to 0.793. A threshold of 0.4 was selected as the primary operating point, as it maximized balance across metrics and minimized false negatives without a substantial loss in specificity.

With a consensus threshold of 0.4, at the patient level, the pipeline achieved an accuracy of 0.834, precision of 0.835, sensitivity of 0.831, specificity of 0.836, F1-score of 0.833, and F3-score of 0.832 (Figure 1a-b). Only one out of 500 individuals (0.2%) failed to receive a prediction because that individual had only one incomplete and uninformative note, which did not produce a valid recommendation output in the desired format. 207 patients were correctly determined by the model to require genetic testing (true positives), 41 were incorrectly thought to need testing when they did not (false positives), 209 were correctly identified as not needing testing (true negatives), and 42 were incorrectly classified as not needing testing when in fact they did (false negatives). Total end-to-end processing time for all 22,510 patient notes on Databricks was 18 hours (Databricks cluster specs: 15.4 LTS, standard NC24ads A100 v4 220 GB, 1 GPU).

Patient-level aggregation of notes significantly improves the overall sensitivity and precision of predictions. The increase in sensitivity from 0.630 (note-level) to 0.831 (patient-level) suggests that aggregating multiple notes enhances diagnostic robustness by mitigating deviations in individual note predictions. This improvement aligns with clinical decision-making processes, where a diagnosis is rarely based on a single clinical entry, which could be incomplete or less informative, but rather on a holistic assessment of multiple records. Furthermore, the F3-score in patient-level evaluation (0.832) highlights the pipeline’s strong performance in prioritizing sensitivity while maintaining diagnostic confidence.

The confidence-based RAG-rerun in our pipeline was motivated by the output probability distribution of the base LLM’s recommendations on a randomly sampled development dataset (50 cases and 50 controls), where misclassifications tend to have a wider probability distribution range (FN and FP in Figure 1e). To address this, we aimed to provide additional relevant information from aortopathy-related literature to help the model correct these errors via RAG. We found that a confidence threshold of 0.5 yields optimal diagnostic performance (Figure 1f), as RAG enhances the LLM’s ability to generate more accurate recommendations for those low-confidence notes. However, applying RAG to high-confidence notes leads to performance degradation. This is likely because the additional information from RAG makes it more challenging for smaller models, such as Llama 3.1-8B-Instruct, to focus on the most relevant clinical details in the patient notes, ultimately hindering the decision-making process.

### Interpretability analysis

Model interpretability is essential in AI-based medical applications as it helps build trust and ensures that the outputs are grounded in meaningful clinical evidence. We built an interpretability pipeline that leverages Captum’s LLM attribution algorithms to enhance the transparency of model predictions.^14^ Figure 2 provides an overview of the interpretability pipeline on a synthetic case example. Both the LLM inputs, including the system prompt and clinical note, and the generated outputs, comprising the genetic testing recommendation and its supporting rationale, are fed into the attribution algorithms to compute the importance scores for input tokens. Various LLM attribution algorithms are supported to analyze the importance of input tokens to the LLM predictions. For example, perturbation-based attribution algorithms systematically ablate individual input tokens - removing or altering them one at a time - and measure the change in output probability to assign an attribution score that reflects each token’s influence. Gradient-based attribution algorithms compute the derivative of the model’s output with respect to each input token embedding to assign importance scores to input tokens.^15^ These scores are then visualized using a blue gradient for the corresponding input tokens, with darker hues indicating a higher impact. Finally, an LLM is used as a filter to retain only medically relevant terms within the patient note, ensuring that the visualization highlights key clinical features for more informed decision-making. More interpretability examples for both synthetic rare disease cases and controls and the required computing time for such analyses are provided in the Supplementary Note 3.

**Figure 2.**
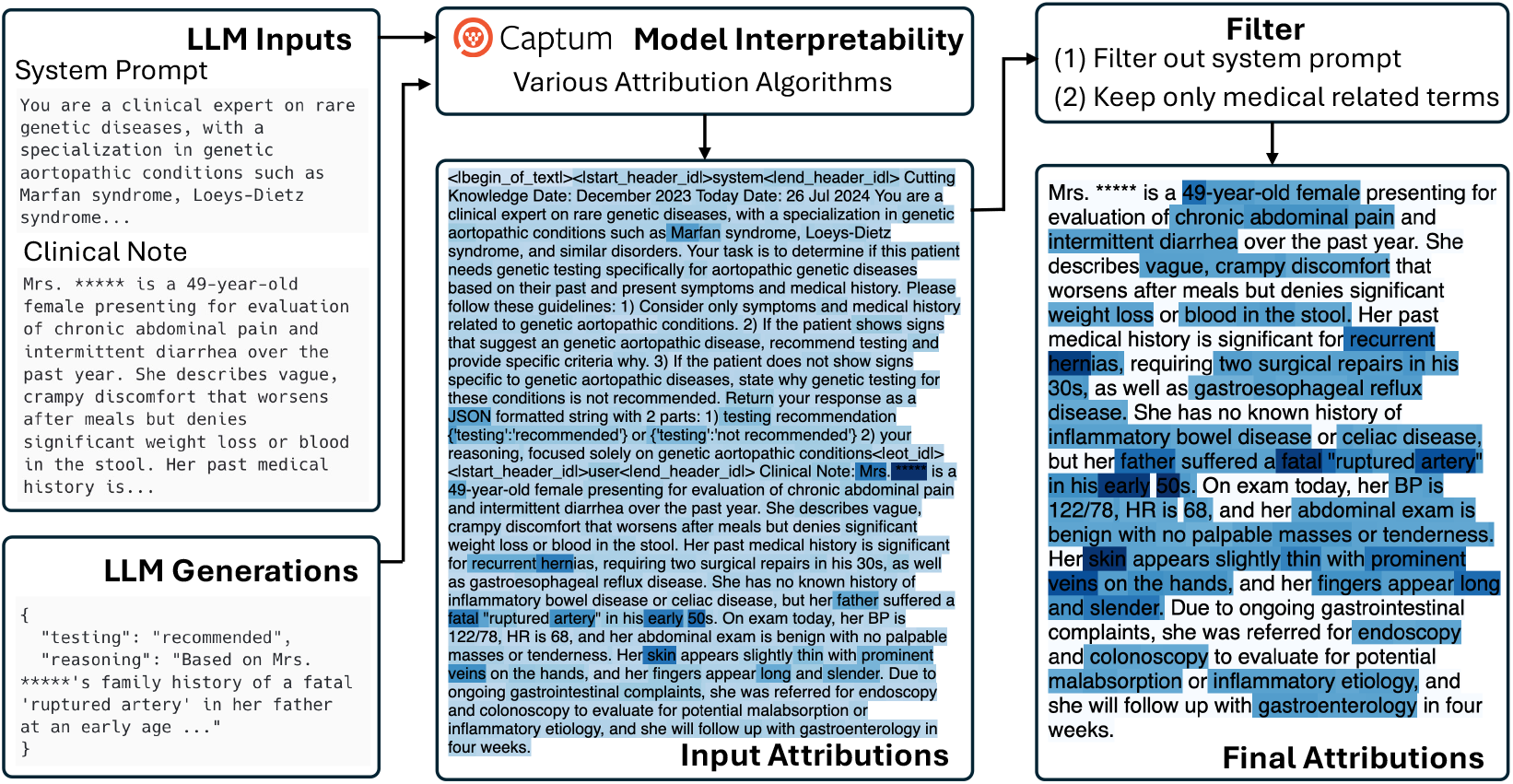
Overview of the interpretability pipeline on a synthetic case example. The full input-output pairs generated by the LLM are processed using Captum’s various LLM attribution algorithms to compute token-level attribution scores, which quantify the contribution of each input token to the model’s output. Another LLM is then employed as a filter to retain only the scores for medically relevant terms, enhancing visualization clarity.

## Discussion

In this study, we present an open-source LLM-enabled framework that provides clinical recommendations for genetic testing for patients with rare aortopathies. While current approaches rely on proprietary models such as GPTs for simplistic medical question-answering setups,^16^ our method prioritizes portability and transparency in evaluating patient clinical notes while providing a framework that could be potentially adapted to other disease domains with appropriate domain-specific adjustment and validation in a privacy-compliant way. Instead of employing RAG in a generic manner without tailoring it to specific clinical scenarios,^17^ our method, employing a RAG-based approach with tailored domain knowledge, provided a genetic testing evaluation for each patient with a 0.834 accuracy and 0.832 F3-score on 22,510 patient notes that did not undergo any content-based preprocessing or filtering step.

Our framework employed a consensus-based aggregation strategy to obtain a holistic assessment for a patient based on the corresponding note-level recommendations. The choice of the consensus threshold represents a clinically meaningful decision that reflects the intended role of the framework in practice. At lower thresholds, the framework prioritizes sensitivity, ensuring that patients with aortopathy-related findings across even a minority of their notes are flagged for review, thus reducing the brisk of missed diagnoses at the expense of a higher false positive burden. At higher thresholds, the model becomes more conservative, improving precision and specificity but increasingly missing patients who may have had aortopathy-related findings documented across only a subset of their clinical notes. In the context of aortopathy screening, where failure to identify a patient for genetic testing can have serious downstream consequences, including undetected risk of aortic dissection and untested at-risk family members, we prioritized minimizing false negatives. A threshold of 0.4 was therefore selected as it achieved near-equivalent sensitivity and specificity, reflecting a balanced operating point that does not disproportionately sacrifice sensitivity for precision. Future prospective validation should confirm whether this threshold generalizes across institutions and patient populations with differing note documentation practices.

Our framework could serve as a **preliminary decision-support tool** to help flag patients who may warrant further clinical evaluation for genetic testing. The traditional model of genetic testing involves the patient first receiving an assessment from their healthcare provider with reasons, a reference to a geneticist, an order for a genetic test, and possibly a reference to a genetic counselor. However, this entire cycle is contingent on the initial provider recognizing the symptoms as being associated with a genetic condition, which are chronically underdiagnosed.^18^ Our approach seeks to provide an automated supplemental recommendation system to the primary care physician that can dynamically assess a patient’s risk as new data from visits is updated.

We tested a variety of parameters in designing this framework, including open-source model types, model parameters such as temperature, prompting text, fine-tuning, RAG corpus chunking and indexing strategies, and other RAG optimizations. We found that varying any of these parameters slightly might have a large impact on performance. For example, RAG performance varies when patient notes are formatted as their original note text compared to when formatted as an extracted list of clinical terms. The format of the desired response specified in the prompt also matters, i.e., requesting a JSON-formatted output produces more consistent results compared to open-ended generations. Furthermore, providing few-shot notes of both cases and controls in the prompt significantly elevates false positive rates in the performance. For these reasons, we selected a reasonable configuration of parameters evaluated on a subset of notes and then tested our framework on a larger set of test notes to assess performance. While future directions for this work may include exploring larger open-source models with diverse architectures, this study focused on performance with portable small models that could easily be integrated into health system computing environments.

In order to provide accessible and relevant insights to a physician, our diagnostic framework also provided reasoning for each genetic testing recommendation. In this study, we were limited in the ability to do a manual review of the 22,510 reasoning outputs; however, we illustrated the utility of attribution algorithms for the interpretability of the responses. Figure 2 shows an example of the attribution algorithm demonstrating how each word in a patient note influences the output prediction. In this case, we see relevant symptoms such as “chronic abdominal pain”, “intermittent diarrhea”, “recurrent hernias”, and “fatal ruptured artery” score highly. This approach could help a physician quickly scan a patient’s history and notes to identify relevant factors that support a diagnostic recommendation by our algorithm. Additional work is needed to validate the relevance of each reasoning in context. Time and computing resources were a limitation of the interpretation analysis (Supplementary Table 2).

This single-institution study demonstrates the feasibility of applying open-source LLMs to longitudinal clinical notes to support identification of patients who may benefit from genetic testing. While the framework was evaluated within one health system and for a specific disease domain, the approach highlights how narrative clinical documentation can be leveraged to assist rare disease screening workflows. Future work will be needed to evaluate performance across institutions, clinical populations, and disease domains. In short, adapting to other disease domains, such as stroke risk or cardiac adverse event, requires tailoring the corpus and prompt accordingly. This paper provides a comprehensive foundation for one to optimize and parameterize for best performance. By demonstrating a transparent and portable framework for LLM-assisted clinical decision support, this work lays the foundation for scalable and privacy-compliant integration of generative AI into routine care, enabling earlier detection and intervention for rare conditions.

Although our work demonstrated promising results, several limitations should be acknowledged. First, the analysis was conducted within a single health system, and documentation practices and referral workflows may differ across institutions, potentially affecting model performance in other clinical environments. Second, physician referral for genetic testing was used as the ground-truth outcome label, as the objective of this study was to model the clinical decision to refer patients for genetic evaluation rather than to predict confirmed molecular diagnoses. While this framing aligns with the intended use of the system as a decision-support tool, referral practices may vary among clinicians and health systems and therefore may partially reflect local clinical behavior. Third, although clinical notes selected for analysis were restricted to those occurring prior to the genetic testing order date, documentation preceding referral may still contain clinical suspicion cues that could influence model predictions. Finally, while the system generates reasoning outputs to support its recommendations, large-scale clinical evaluation of these reasoning explanations was beyond the scope of this study and warrants further investigation.

## Methods

We developed an LLM-enabled recommendation pipeline to flag patients who would benefit from genetic testing for rare genetic diseases and provide corresponding reasoning. We utilize techniques such as retrieval augmented generation (RAG) and fine-tuning on a curated genetic aortopathy-related corpus to inject relevant medical knowledge into the base LLM to bolster performance in providing more reasonable and accurate recommendations.

### Patient data

Clinical progress notes from patients in Penn Medicine’s health system between 2019 and 2024 were utilized to develop and test the pipeline. The **case cohort** consisted of 273 patients who were referred by a physician for genetic testing for a suspected connective tissue disorder associated with genetic aortopathy. In this study, physician referral for genetic testing was used as the operational ground-truth label, reflecting real-world clinical decision-making regarding suspected genetic disease rather than confirmed molecular diagnoses. This framing reflects the intended use case of the system as a decision-support tool for identifying patients who may benefit from genetic testing. Progress notes style and template (or lack there of) varies across Penn Medicine based on department, physician, and date. For each case patient, progress notes from all encounters occurring within the **12 months prior to the genetic testing order date** were included in the analysis. Notes written after the referral order were excluded to ensure that model predictions were based on clinical documentation preceding the testing decision. The **control cohort** was drawn from the Penn Medicine BioBank^19^ and consisted of 1,200 individuals matched to the case cohort by age and sex. Control patients had **no documented referral for genetic testing and no predicted loss-of-function variants in genes** associated with genetic aortopathies based on available sequencing data (Supplementary Table 1). Controls were not preselected or filtered for any other clinical criteria so as to preserve the naturally occurring distribution of phenotypes found, including other cardiac conditions. All available clinical progress notes for control patients were included to represent routine clinical documentation in individuals without suspected aortopathy. From these pools, a **development cohort** consisting of 50 cases and 50 controls was randomly sampled for parameter selection and pipeline configuration. The final **testing cohort** consisted of 250 cases and 250 controls was randomly sampled from the pool, comprising 22,510 total clinical notes. Figure 3 illustrates the statistics for the testing patient cohort.

**Figure 3.**
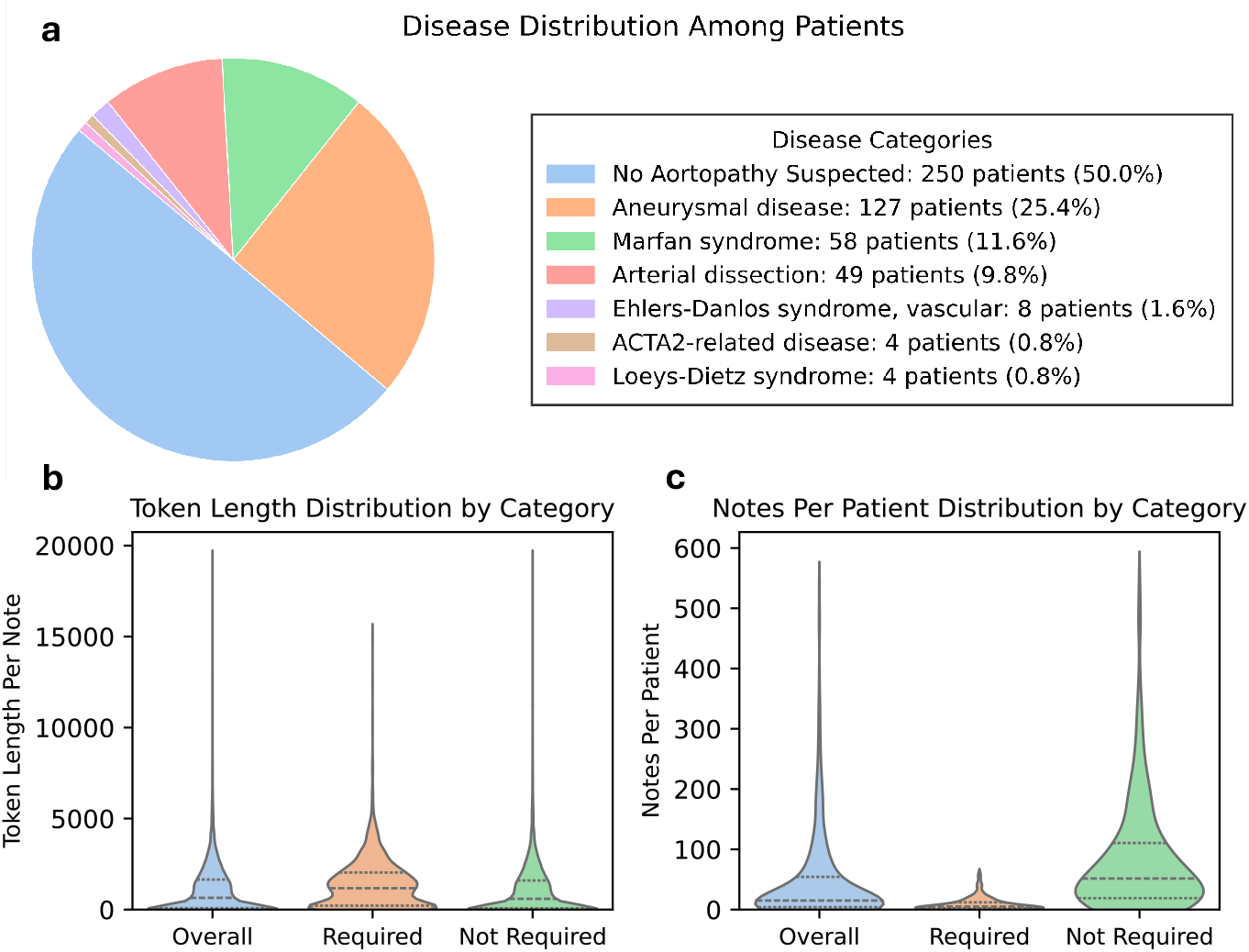
Clinical note statistics for the randomly sampled testing patient cohort. **a** Disease distributions of the 500 individuals within the randomly sampled case/control testing cohort. **b** The violin plot showing token length distributions for the patient notes by category: all patient notes, notes of patient required testing (cases), and notes of patients not required testing (controls). **c** The violin plot showing distributions of the total number of notes per patient by category.

### Model selection

In the healthcare domain, data privacy and regulatory constraints impose significant limitations on model selection. Medical institutions usually have strict policies governing data access, making reliance on proprietary models like ChatGPT, which are challenging for local deployment and often require external API calls for inference, less viable despite their strong performance. To ensure data privacy and enable on-premise deployment, we opted for open-source models that can be seamlessly deployed within a local secure computing environment without external dependencies. To determine the suitable base model for this study, we evaluated several popular open-source models, including Llama 2-7B-Chat,^20^ Mistral-7B-Instruct,^21^ Llama 3-8B-Instruct, Llama 3.1-8B-Instruct, and Llama 3.1-70B-Instruct.^12^ We compared these models on the development dataset with 50 cases and 50 controls and evaluated their performance at both the note and patient levels. Our analysis revealed that Llama 3 and Llama 3.1 models consistently outperformed the others (Figure 4a-c). In contrast, Llama 2-7B-Chat and Mistral 7B-Instruct-v0.3 exhibited an overly conservative bias, mis-classifying most patients as positive (Figure 4c). This high false positive rate makes them unsuitable for clinical applications.

**Figure 4.**
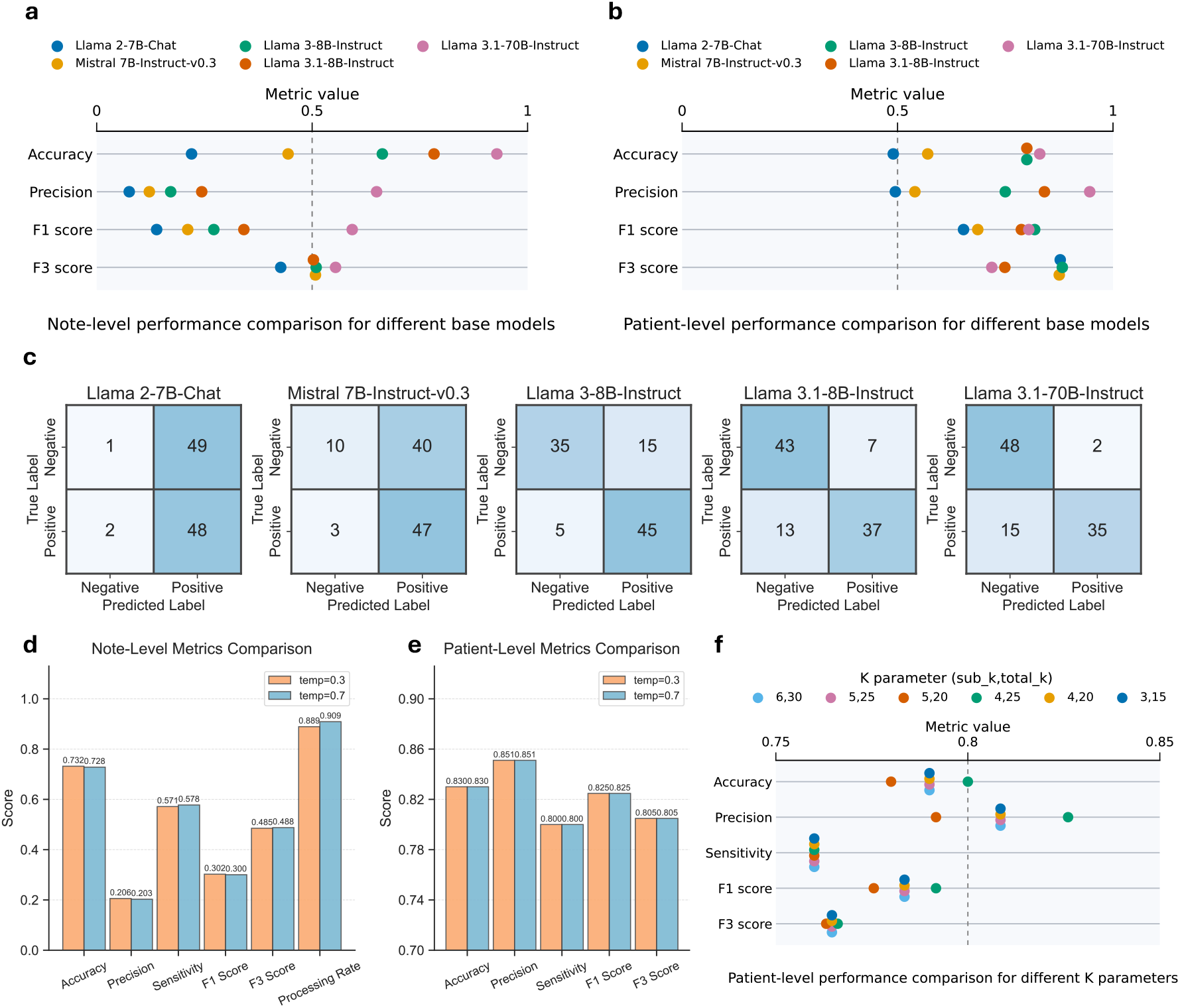
Performance comparisons for various pipeline settings. **a** Note-level performance comparison across multiple LLMs on the development dataset with 50 cases and 50 controls. **b** Patient-level performance comparison across multiple LLMs on the development dataset. **c** Patient-level confusion matrices for multiple LLMs on the development dataset. **D** Note-level performance comparison between temperatures 0.3 and 0.7 on the development dataset. **e** Patient-level performance comparison between temperatures 0.3 and 0.7 on the development dataset. **f** Patient-level performance comparison among different K parameters in RAG on the development dataset.

In addition to performance on the development dataset, another crucial factor in our model selection was the context window length of the LLM, which refers to the maximum number of tokens an LLM can process before collapsing and generating uncontrollable content. Llama 3.1 models support a 128K-token context window, while Llama 3 models are limited to 8K tokens. In our datasets, some patient notes exceed 8K tokens (Figure 3b), making a longer context window essential for accurate analysis. Additionally, techniques such as RAG used in our pipeline introduce additional tokens to the LLM. The 128K-token context window of Llama 3.1 allows for full integration of the lengthy patient notes as well as the retrieved knowledge. Considering these factors, although Llama 3 demonstrated strong performance on the development set even with lower false negative rates, we opted for Llama 3.1 due to its substantially larger context window, which enables robust handling of longer clinical notes and provides greater flexibility for incorporating additional context through methods such as RAG.

When comparing 8B and 70B variants of Llama 3.1, Llama 3.1-70B-Instruct generally has stronger performance across many metrics but is significantly less computationally efficient. Inference with the 70B model requires around 140GB of VRAM, typically necessitating a compute node with four to eight GPUs. In contrast, Llama 3.1-8B-Instruct is much more computationally efficient and capable of running on a single 24GB GPU, making it feasible and accessible for a broader range of medical institutions. Furthermore, at the patient level, the 8B model outperformed the 70B model in sensitivity and F3-score, two important metrics in rare disease identification tasks. Considering these factors, we finally selected Llama 3.1-8B-Instruct as the base LLM in our study to strike a balance between data privacy, computational efficiency, and clinical relevance, ensuring scalability and accessibility for real-world medical usage. Additionally, if resources permit, the base model can be seamlessly upgraded to a larger and more powerful variant within the same pipeline.

### LLM temperature selection

Temperature is a crucial hyperparameter in LLM inference, particularly in medical applications: a lower temperature encourages more deterministic and stable outputs, which is desirable for clinical reasoning, while a slightly higher temperature introduces variability that may enhance flexibility. Therefore, it is essential to identify a temperature setting that provides sufficient diversity in outputs without compromising reliability. Given these considerations, we conducted experiments using two temperature values: 0.3 and 0.7, aiming to assess their impact on pipeline performance. The choice of 0.3 and 0.7 was guided by the need to maintain stability while allowing a degree of flexibility. A temperature lower than 0.3 would result in overly rigid and deterministic responses, potentially limiting the model’s ability to generalize across diverse clinical narratives. Conversely, a temperature significantly higher than 0.7 could introduce excessive randomness, leading to inconsistencies in medical reasoning and factual accuracy.

For evaluation, we tested our pipeline on the development dataset. We observed that at the note level, temperature 0.7 outperformed 0.3, while at the patient level, both settings produced comparable results (Figure 4d-e). Beyond performance accuracy, we also analyzed the number of successfully processed notes. We defined a failed note processing instance as any output that did not conform to the expected structured format and was therefore unparseable. The results showed that temperature 0.7 successfully processed 4,080 notes, whereas temperature 0.3 processed 3,991 notes, resulting in an additional note processing ratio of 2.2% for temperature 0.7. We hypothesize that this is due to the model’s increased flexibility at 0.7, which allows it to escape rigid output patterns that could contribute to systematic parsing failures. In contrast, at a lower temperature (0.3), the model tends to generate more deterministic and repetitive outputs, making it susceptible to strict formatting constraints and increasing the likelihood of failure when slight variations occur in input structures. The added variability at 0.7 may have helped prevent repeated failure patterns, ultimately improving output consistency across different cases. Given these findings, we selected temperature 0.7 as the final setting for our pipeline. This decision was based on its ability to maintain comparable performance at the patient level while improving processing efficiency by reducing formatting-related failures. This balance between reliability and robustness is critical for deploying LLMs in clinical settings, ensuring both interpretability and operational efficiency in real-world applications.

### Knowledgebase curation

To improve the capability of general-purpose LLMs in aortopathy-related domains for more accurate recommendations, we curated a specialized medical knowledgebase for injecting domain-specific knowledge into the models. This knowledgebase includes full-text PubMed Central papers on genetic aortopathies published within the last 5 years. We used the following keywords to select a scope of papers, and then selected further based on journals and authors of interest: [“marfan”, “aneurysm”, “cardi”, “ehlers”, “danos”, “erdheim”,“chester”, “thoracic”, “taad”, “loeys”, “dietz”, “sotos”, “vascular”, “shprintzen”, “goldberg”, “dissection”, “aort”, “syndrome”, “symptoms”]. NCBI’s clinical guidelines for genetic diseases, GeneReviews, were also included, in addition to relevant aortopathy-related medical textbooks. We leveraged MinerU,^22^ a state-of-the-art PDF parsing tool, to accurately convert PDF files into raw text files. This process resulted in a comprehensive yet compact rare genetic aortopathy knowledge corpus comprising a total of 1,064 PDF documents.

### Retrieval augmented generation

The curated knowledgebase can enhance the performance of LLM through retrieval-augmented generation (RAG). RAG retrieves relevant text chunks from the knowledgebase based on the patient’s clinical notes and appends them to the input of the LLM. The additional contextual information helps improve the LLM’s ability to generate more accurate predictions. Efficient retrieval in RAG is essential for LLM performance improvement; however, standard text-chunking and indexing strategies often fail to maintain semantic coherence in medical texts.

To address this, we adopted a strategy inspired by MedRAG,^23^, which combines semantic-aware chunking for the knowledgebase with multi-retriever fusion during indexing. Standard chunking methods, which split long texts into chunks with a certain number of characters or words, often disrupt the contextual integrity of the split chunks, thus reducing retrieval performance. Therefore, we implemented a semantic double-pass merging (SDPM) approach: first grouping semantically similar sentences, then merging adjacent similar chunks to preserve context and minimize redundancy. In our SDPM, we utilized a similarity threshold of 0.5, a chunk size of 512 tokens, and a minimum of one sentence per chunk. In the second merge pass, we applied a skip window size of one, allowing the algorithm to bypass a single intervening chunk when evaluating semantic similarity. This ensured that each final chunk encapsulates a complete and meaningful unit of information.

For indexing, we adopted a hybrid retrieval system that integrates both lexical and dense retrieval models. BM25 was used for lexical matching,^24^ enabling precise keyword-based search, while the domain-specific embedding model MedCPT^25^ facilitated semantic retrieval based on the embedding similarity. These two methods were combined using reciprocal rank fusion to balance lexical precision with semantic depth.^26^ Additionally, given that clinical notes often exceed the 512-token limit of the MedCPT embedding model, long clinical notes were segmented into smaller, context-preserving chunks, and each of which was retrieved independently. The retrieved segments were then aggregated to maintain comprehensive and relevant contextual information. We used a token-based chunking strategy for the clinical notes, which splits input text into segments based on a predefined token limit and overlap. This ensures readability and keeps words intact, while also minimizing the number of sub-notes, thereby reducing redundant retrieval. Each sub-note chunk has a fixed length of 512 tokens with a 25-token overlap to maintain contextual continuity across chunks. To improve retrieval performance, we introduce two additional parameters: (1) sub_k, the number of retrieved chunks per sub-note chunk, and (2) total_k, the total number of retrieved chunks across all sub-note chunks. We chose these parameters through experiments on the development cohort with 50 cases and 50 controls. We found that setting sub_k = 4 and total_k = 25 strikes the best balance between sensitivity and precision (Figure 4f).

### Model fine-tuning

Fine-tuning is also considered effective to inject domain-specific knowledge into LLMs.^27^ Traditional fine-tuning typically refers to supervised fine-tuning, where labeled question-answer pairs are used to update the LLM for better performance on specific tasks. However, in this task, we required the LLM to generate both genetic testing recommendations and corresponding reasoning to aid physicians in decision-making. Collecting a sufficiently large, expert-annotated notes training set for this purpose is challenging, and furthermore, training the model on notes limits generalizability when evaluating new patient histories and symptoms. In other words, we did not want to limit the model’s exposure to only a certain set of patient profiles through training on notes. Therefore, we opted for continual pre-training on the collected unlabeled aortopathy corpus (the curated medical knowledgebase mentioned before) to enhance the model’s understanding of genetic aortopathy. We employed Low-Rank Adaptation (LoRA), a parameter-efficient tuning method proven effective in low-data scenarios.^28^ However, our tests on the development dataset indicated that such continual pre-training did not improve the LLM’s performance in our tasks, likely due to limited data size and catastrophic forgetting. Consequently, fine-tuned models were not utilized in our final pipeline. Further details and explanations are provided in the Supplementary Note 2.

### Pipeline Implementation

The pipeline first produces predictions at the note-level and then the patient-level with two main components: (a) a recommendation for genetic testing (“recommended” or “not recommended”), and (b) a corresponding reasoning for each prediction. The pipeline is composed of the following main steps:

i. LLM recommendation for each patient note (substeps 1 and 2 in Figure 5);
ii. Confidence evaluation for the recommendation (substep 3 in Figure 5);
iii. RAG-based re-inference for low-confidence notes with retrieved relevant domain knowledge (substeps 4, 5, and 6 in Figure 5);
iv. Consensus-based aggregation of note-level recommendations to generate final patient-level recommendations (substeps 7 and 8 in Figure 5);

**Figure 5.**
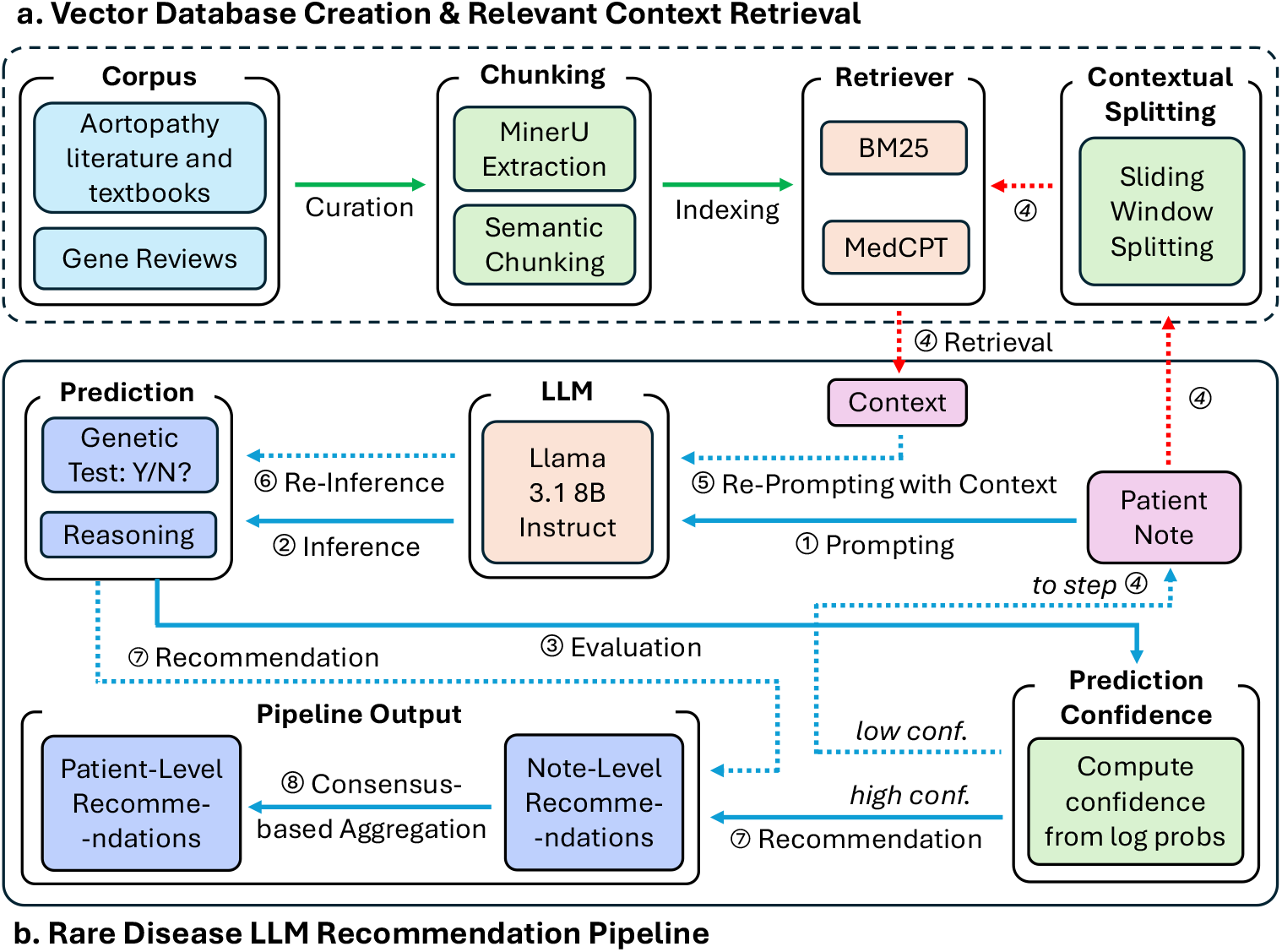
Overview of the LLM-enabled genetic testing recommendation pipeline for rare genetic aortopathies. **a** The raw aortopathy corpora are exacted via MinerU and split into semantically meaningful chunks, which are then indexed to vector databases for further retrieval. **b** Each patient note is passed to a Llama 3.1-8B-Instruct model for generating genetic testing recommendations, with the generation confidence evaluated. Low-confidence notes retrieve relevant contextual information from the vector database for relevant context and re-prompt the LLM for a final note-level recommendation. All note-level recommendations for an individual patient will be evaluated using a consensus-based aggregation approach to generate the final patient-level recommendation for genetic testing.

In step (i), each clinical note for an individual is passed to the LLM independently for inference to generate predictions composed of two parts as JSON-formatted strings: a binary classification and an explanatory reasoning output. No pre-processing was done on the clinical notes, though the pipeline is modular in that one could pass notes through filters if desired. Given stochasticity in response generation, the number of rerun iterations for a poorly-formatted or null-response note can be set (default = 3). Responses for notes that do not meet the desired format will be dropped after several rerun iterations. To enhance the recommendation reliability, in step (ii), a confidence score is derived from the log probability of the tokens for “recommended” or “not recommended” in the model output. If the probability is below a certain threshold, the model response is considered uncertain, and a RAG-based re-inference will be triggered by incorporating relevant medical knowledge retrieved from the curated database (step iii). This confidence-based RAG-rerun approach was informed by an initial run on the development cohort with 50 cases and 50 controls, which demonstrated that false predictions tend to have lower confidence values, and RAG can improve LLM’s performance on low-confidence notes (Figure 1c-d). To derive a final patient-level recommendation, in step (iv), we apply a consensus-based aggregation strategy across all available notes for each patient. If more than 50% of a patient’s clinical notes are classified as “recommended”, the final output is set to “recommended”. This approach reduces the impact of uninformative and incomplete individual notes while preserving sensitivity to relevant diagnostic signals, and mitigates noise introduced by clinical visits for concerns unrelated to a potential suspected aortopathy. The final system output includes both the recommendation and a reasoning summary to bolster clinician interpretation.

The pipeline is designed to be adaptable across healthcare systems by adjusting the base model, prompt, retrieval database, and confidence threshold. When deploying on a new task, parameters can be tuned iteratively to optimize performance before full-scale implementation. Additionally, as the pipeline is used, high-quality cases can also be incorporated into the retrieval database, progressively enhancing its performance.

### Prompting

Prompt is crucial for LLM performance as it defines the context, scope, and intent of the output. Our prompt consists of general guidelines outlining the scope of the evaluation, clinical task of recommendation and reasoning, and output format specifications as a JSON-formatted string for easy and automated result parsing (Figure 6).

**Figure 6.**
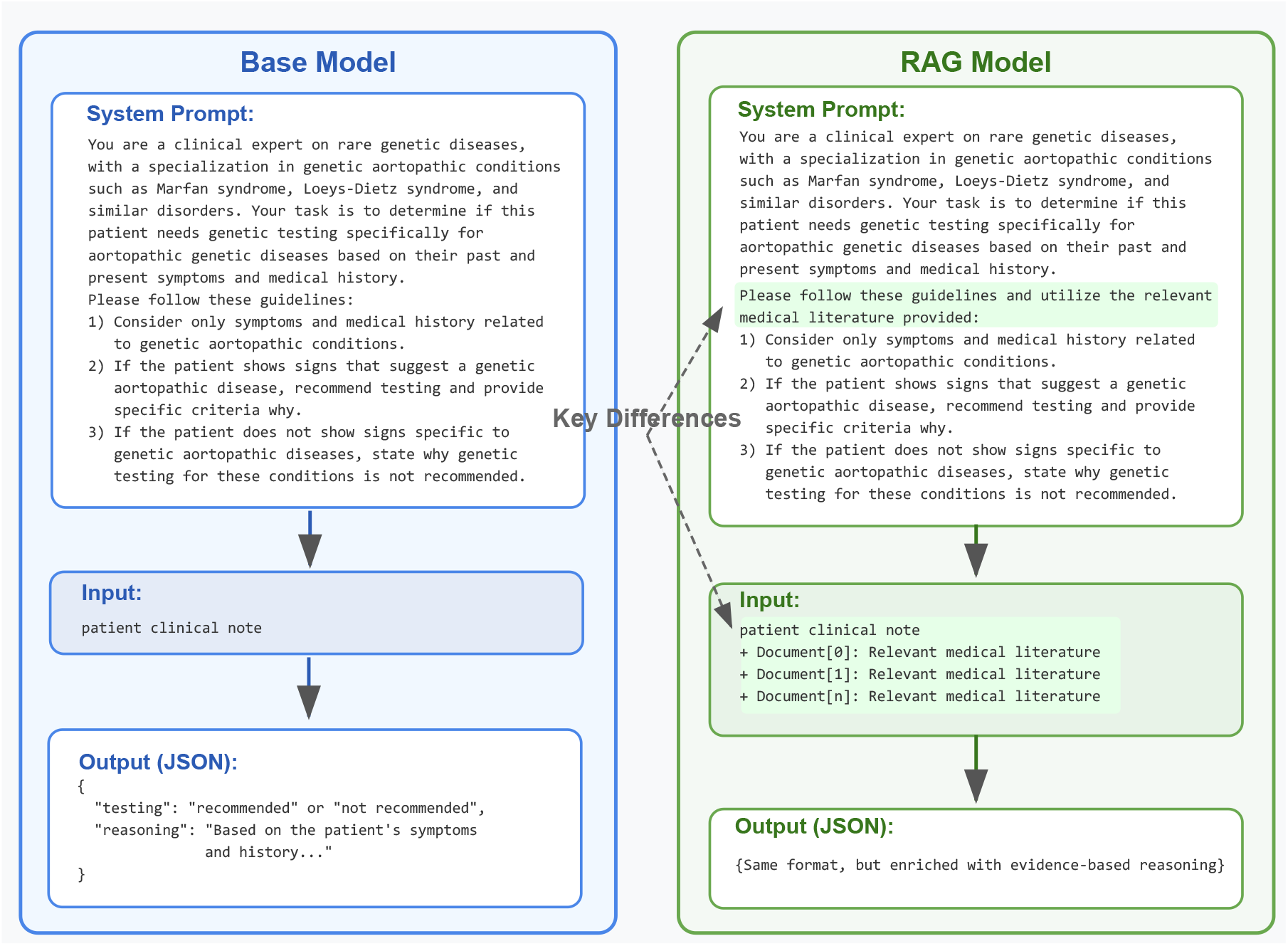
Prompting schematic. In the base LLM inference, the left prompt is utilized. Upon inference with RAG on low-confidence notes, the right prompt, together with the retrieved relevant knowledge are provided to the LLM.

## Supporting information

All Supplementals

## Data availability

Patient notes data used in this study will not be made available to anyone due to HIPAA compliance and privacy restrictions for Penn Medicine. Our vector database capturing the curated aortopathy knowledgebase will be released on GitHub upon publication to facilitate reproducibility and reuse.

## Code availability

Implementation code and documentation for the pipeline will be released on GitHub upon publication to facilitate reproducibility and reuse.

## Acknowledgements

P.S. is supported by Genomic Medicine T32 Training Grant 5T32HG009495-08. This research uses resources of the Argonne Leadership Computing Facility, a U.S. Department of Energy (DOE) Office of Science user facility at Argonne National Laboratory and is based on research supported by the U.S. DOE Office of Science-Advanced Scientific Computing Research Program, under Contract No. DE-AC02-06CH11357. This research also utilizes computing resources provided by the National Artificial Intelligence Research Resource (NAIRR) Pilot, supported by award NAIRR240008.

## Author contributions

P.S., Z.L., and Z.Y. conceptually designed the study, conducted the analysis, and drafted the manuscript. T.N., A.R., V.K., G.S., R.P., T.D., R.M., and A.V. provided guidance and feedback in study design and analysis. P.S., C.M., Z.R., and A.V. had full access to all the data in the study and were responsible for the integrity and accuracy of the data analysis. C.M. and Z.R. deidentified the clinical data and P.S. and A.V. accessed and utilized the data. All authors critically edited, read, and approved the final manuscript. P.S., Z.L., and Z.Y. contributed equally and shared co-first authorship. R.M. and A.V. jointly supervised this work.

