## Supplementary material for "Leveraging Open-Source Large Language Models to Identify Undiagnosed Patients with Rare Genetic Aortopathies": All Supplementals

### Supplementary Note 1: Genes and associated diseases

Supplementary Table 1 shows a list of genetic aortopathy-related genes and their associated diseases.

| Genes | Disease | Genes | Disease | Genes | Disease |
|---|---|---|---|---|---|
| *ACTA2* | Familial TAAD | *FBN2* | Marfan | *PLOD1* | Ehlers-Danlos |
| *ACTA2* | Arterial dissection | *FKBP14* | Marfan | *PRKG1* | TAAD |
| *ACTA2* | Marfan | *FLNA* | Ehlers-Danlos | *SLC2A10* | Arterial Tortuosity Syndrome |
| *ADAMTS2* | Ehlers-Danlos | *FOXE3* | Familial TAAD | *SMAD2* | Loeys-Dietz |
| *ASH2A* | Marfan | *FOXE3* | TAAD | *SMAD2* | TAAD |
| *B4GALT7* | Ehlers-Danlos | *HCN4* | TAAD | *SMAD3* | Loeys-Dietz |
| *C1R* | Ehlers-Danlos | *IDS* | MPSI | *SMAD4* | TAAD |
| *CBS* | Arterial dissection | *IDUA* | Scheie syndrome | *SMAD6* | TAAD |
| *CHST14* | Ehlers-Danlos | *IQSEC2* | IQSEC2-related | *SRY* | Klinefelter |
| *COL1A2* | Ehlers-Danlos | *KCTD10* | KCTD10-related | *TGFB2* | Loeys-Dietz |
| *COL3A1* | Ehlers-Danlos | *LOX* | TAAD | *TGFB3* | Loeys-Dietz |
| *COL5A1* | Ehlers-Danlos | *MED12* | TAAD | *TGFBR1* | Loeys-Dietz |
| *COL5A1* | MASS | *MYH11* | Familial TAAD | *TGFBR2* | Loeys-Dietz |
| *COL5A2* | Ehlers-Danlos | *MYLK* | Familial TAAD | *TNXB* | Ehlers-Danlos |
| *EFEMP2* | TAAD | *NOTCH1* | TAAD | *TNXB* | TAAD |
| *FBN1* | Marfan | *NSD1* | Sotos | *ZNF469* | TAAD |
| *FBN1* | MASS | *PKD1* | TAAD | | |

**Supplementary Table 1. List of genes and associated diseases**

### Supplementary Note 2: Details on model fine-tuning

To fine-tune the pre-trained LLMs, we utilized the unlabeled, curated aortopathy-related knowledgebase and applied next-token prediction for model continual pretraining in an unsupervised manner, aiming to inject domain-specific knowledge without requiring labeled training data. Given the relatively small size of the training dataset, we employed Low-Rank Adaptation (LoRA), a parameter-efficient tuning method proven effective in low-data scenarios by updating only a small set of model adapters. We trained both small and large LoRA adapters for Llama 3.1-8B-Instruct, with the following training configurations: (1) LoRA-small: rank of 16 and scaling factor of 32, applied only to the Q and V matrices of the LLM; (2) LoRA-large: rank of 32 and scaling factor of 64, applied to all matrices within the LLM. The models were trained using the AdamW optimizer[1] with a learning rate of $2 \times 10^{-5}$, a weight decay factor of 0.01, and a learning rate decay factor of 0.85. The training batch size is set to four, with each batch containing 8192 tokens. The model was fine-tuned for one epoch on the entire curated corpus using one NVIDIA A100 GPU, with training taking approximately 20 hours.

The fine-tuned models were evaluated on their ability to correctly recommend patients for genetic testing on a small development dataset of 25 cases and 25 controls, compared to the base Llama 3.1-8B-Instruct model. Results indicated that both fine-tuned models became significantly more conservative, classifying all 50 individuals as requiring genetic testing (Supplementary Figure 1). This resulted in a patient-level accuracy of 0.5, precision of 0.5, and sensitivity of 1.0. Several key challenges might contribute to this suboptimal performance, including the limited availability of suitable training data and the difficulty of fine-tuning pre-trained LLMs without compromising their general knowledge understanding and reasoning capabilities.

For the training data, though several medical Q&A and conversation datasets are available,[2,3] they primarily consist of general Q&A pairs or patient-doctor dialogues for tuning medical language models on broad medical questions. These datasets are not suitable for tuning an LLM to address highly specific and narrow tasks, such as recommending genetic testing for patients with step-by-step reasoning, as in our study. Therefore, supervised instruction tuning of pre-trained LLMs requires a substantial amount of labeled Q&A pairs tailored to the relevant applications, but manually collecting such labeled training data from domain experts is extremely costly and time-consuming, making it highly impractical for the use case in this study. Alternatively, automatically generating labeled data using the pre-trained LLMs or advanced closed-source models like GPT has proven useful for building models that better follow instructions,[4] and provide improved answers to general domain-specific questions.[5] However, these automatically generated datasets may still fail to capture the intricacies of highly specialized tasks, underscoring the need for more targeted data collection methods. Moreover, a bias may be introduced if the same model used to generate the data is used for an inference task on the data. Given the scarcity of suitable instruction-tuning data, unsupervised fine-tuning (i.e., continual pre-training) remains a more viable option for most highly specific medical applications, as we did in this case study. Unsupervised fine-tuning only requires an unlabeled relevant corpus, such as textbooks or research articles, and implicitly injects domain knowledge by exposing the model to domain-specific corpora through next-token prediction tasks.

While data collection is easier for unsupervised fine-tuning, the tuning process itself is complicated. It is not as simple as updating model parameters using domain-specific datasets, as this can result in catastrophic forgetting, where previously learned pre-trained knowledge is lost, or it may impair the model's instruction following and knowledge understanding abilities, as observed in our results. Effective fine-tuning requires complex data curation, source selection, and a well-calibrated learning rate schedule,[6] which is even more challenging given the opacity of pre-training details for many state-of-the-art open-source LLMs. Moreover, as LLMs incorporate increasingly larger datasets (e.g., Llama 3.1 models are pre-trained on over 15 trillion tokens), there is a high likelihood of overlap between the fine-tuning and pre-training corpora, particularly when fine-tuning datasets are sourced from publicly available materials, which may further reduce the effectiveness of fine-tuning.

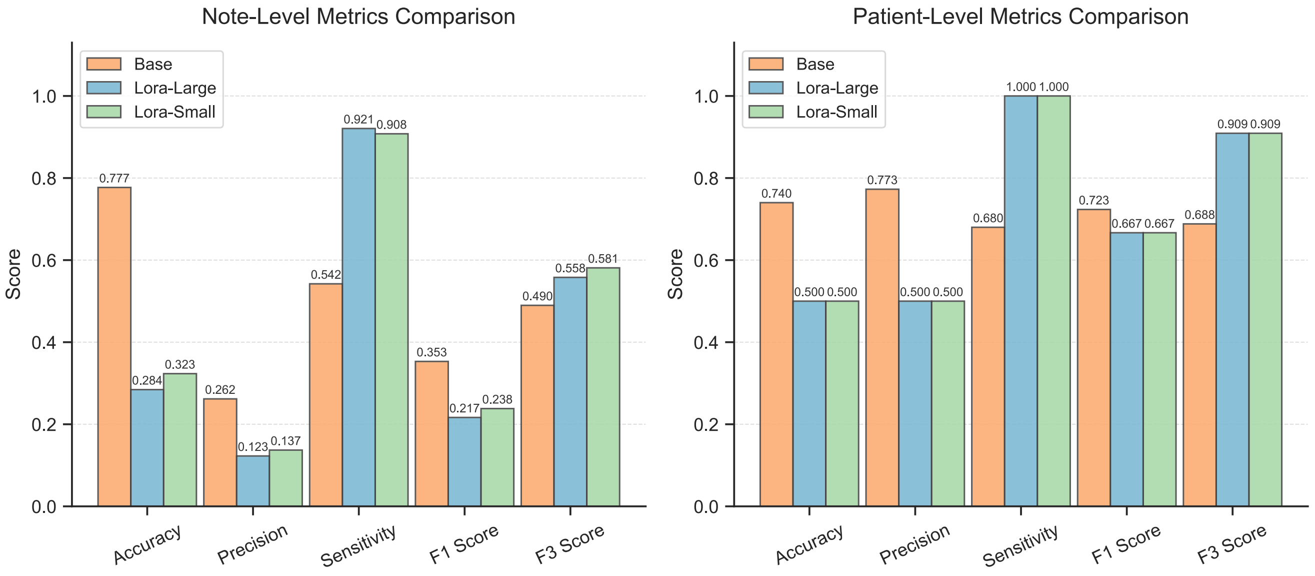


**Supplementary Figure 1.** Model performance comparison between Llama 3.1-8B-Instruct base model and two fine-tuned versions.

### Supplementary Note 3: Additional model interpretability examples

We examined various attribution algorithms provided by Captum: the perturbation-based algorithm, integrated gradients with 50 and 10 approximation steps, and the gradient x activation. Supplementary Figure 2 shows the model interpretability outputs for three synthetic case samples and three synthetic control samples using these algorithms. It is important to note that these attribution methods vary in their computational demands and hardware requirements. Detailed comparisons of average runtime and GPU usage are provided in the Supplementary Table 2, where the average GPU time is equal to the multiplication of the number of required GPUs and the average computing time.

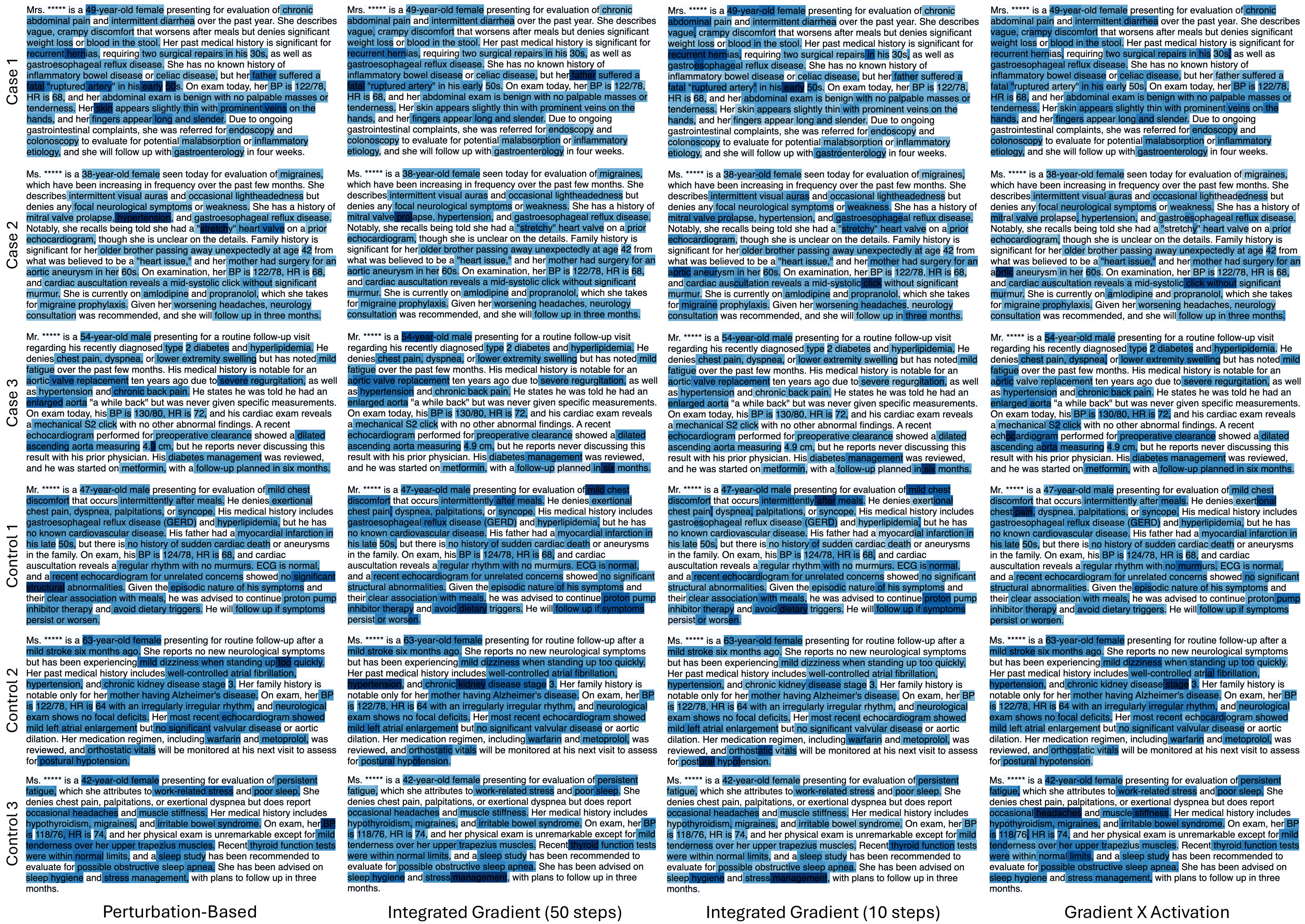


**Supplementary Figure 2.** Additional model interpretability examples on three synthetic case samples and three synthetic control samples.

| Algorithm | # of NVIDIA A100 GPUs | Avg. Computing Time (min) | Avg. GPU Time (min) |
| --- | --- | --- | --- |
| Perturbation-Based | 1 | 152.28 | 152.28 |
| Integrated Gradients (50 steps) | 8 | 20.17 | 161.36 |
| Integrated Gradients (10 steps) | 8 | 4.75 | 38.00 |
| Gradient X Activation | 1 | 1.13 | 1.13 |

**Supplementary Table 2.** Average computation time and number of NVIDIA A100 GPUs required for various LLM attribution algorithms.
